# Neural Tube Defects in the Czech Republic: Incidence, Prenatal Diagnosis and Sex Distribution

**DOI:** 10.64898/2026.01.26.26344821

**Authors:** Antonín Šípek, Natálie Grošup-Friedová, Marek Malý, Jan Klaschka, Antonín Šípek

**Author notes:** **Corresponding author:** Antonín Šípek, Jr., Institute of Biology and Medical Genetics, General University Hospital and First Faculty of Medicine, Charles University, Albertov 4, 128 00, Prague 2, Czech Republic.

## Abstract

Neural tube defects (NTDs) represent frequent and severe congenital anomalies of the central nervous system, including anencephaly, spina bifida, and encephalocele. This retrospective study evaluates the occurrence, prenatal diagnosis, and sex distribution of NTDs in the Czech Republic during the period 1961–2020. Data were obtained from the National Registry of Congenital Anomalies within the National Registry of Reproductive Health. Both prenatally and postnatally diagnosed cases of anencephaly, spina bifida, and encephalocele were analyzed. A total of 2,521 cases of anencephaly, 3,391 cases of spina bifida, and 704 cases of encephalocele were recorded. Prenatal diagnosis with subsequent termination of pregnancy accounted for a substantial proportion of cases, particularly in anencephaly. The mean total incidence per 10,000 live births was 2.91 for anencephaly, 4.38 for spina bifida, and 1.24 for encephalocele. Sex distribution analysis across six consecutive decades demonstrated a persistent predominance of affected females in spina bifida, with statistically significant differences in selected periods. In contrast, anencephaly and encephalocele showed a female predominance only in earlier decades, whereas a higher proportion of affected males has been observed in recent years. Although advances in prenatal diagnostics have markedly reduced the incidence of NTDs among live-born children, the overall population incidence of these defects has remained stable.

## Introduction

Neural tube defects (NTDs) constitute a major group of congenital malformations of the central nervous system resulting from failure of neural tube closure during early embryogenesis (Avagliano et al., 2018). According to standard classification, NTDs are divided into three principal categories: anencephaly, spina bifida, and encephalocele. Anencephaly represents the most severe form and is characterized by absence of the cranial vault and major parts of the brain due to failure of closure of the rostral neuropore, typically occurring between the 23rd and 26th day after conception. This defect is lethal, with most affected pregnancies ending in spontaneous abortion, stillbirth, or early neonatal death (Avagliano et al., 2018; Salari et al., 2022).

The etiology of anencephaly, as well as other NTDs, is complex and largely multifactorial. Most cases are sporadic, although familial recurrence has been reported, suggesting a genetic contribution in a minority of cases. Both autosomal recessive forms and polygenic inheritance patterns have been described (Record and McKeown, 1950; Iffy, 1963). Environmental factors also play an important role, with maternal folate deficiency being one of the most consistently identified risk factors. Adequate periconceptional folic acid intake has been shown to significantly reduce the risk of NTDs, including anencephaly and spina bifida (Morris et al., 2021).

Spina bifida arises from incomplete closure of the neural tube, most commonly affecting the lumbosacral region, although any spinal level may be involved (Ata et al., 2016). Depending on severity, spina bifida is classified into spina bifida occulta, meningocele, and myelomeningocele, the latter being the most severe form associated with substantial neurological morbidity. Similar to anencephaly, spina bifida is considered a multifactorial condition influenced by genetic susceptibility and environmental exposures (Hassan et al., 2022). Despite this, approximately 95% of affected children are born to parents without a positive family history. Increased risk has been reported in pregnancies complicated by maternal diabetes, epilepsy, obesity, hyperthermia, or exposure to certain antiepileptic drugs. Several studies have also reported a higher prevalence of spina bifida among females compared to males (Copp et all., 2015).

Encephalocele is the least frequent of the three major NTDs and is characterized by herniation of brain tissue and meninges through a defect in the skull, most commonly in the occipital region. It may occur as an isolated defect or as part of complex genetic syndromes, such as Meckel–Gruber syndrome (Hartill et al., 2017). Encephalocele is often associated with additional central nervous system anomalies, including hydrocephalus. The sex distribution of encephalocele, similarly to other NTDs, has been reported to vary across populations and time periods (Protzenko et al., 2021; Akyol et al., 2022).

Although advances in prenatal diagnostics have substantially reduced the number of live-born children affected by NTDs, the overall population incidence has not declined in many European countries (Morris et al., 2021). Moreover, sex-specific differences in the occurrence of NTDs remain insufficiently explained (Brook et al., 1994; Deak et al., 2008). The present study aims to analyze long-term trends in incidence, prenatal diagnosis, and sex distribution of anencephaly, spina bifida, and encephalocele in the Czech Republic over a 60-year period.

## Materials and Methods

Data for this retrospective population-based study were obtained from the National Registry of Congenital Anomalies, which is part of the National Registry of Reproductive Health maintained by the Institute of Health Information and Statistics of the Czech Republic. Additional data on prenatal diagnostics were derived from records of the Czech Society of Medical Genetics for the period 1985–2020. The registration process of congenital anomalies in the Czech Republic is population-wide and compulsory by the Czech national law (Šípek et al., 2014)

Cases of neural tube defects included both live-born children and prenatally diagnosed pregnancies terminated due to NTDs. Eligible cases were identified according to the International Classification of Diseases, 10th Revision (ICD-10), using the following diagnostic codes: anencephaly (Q00 or Q00.0), spina bifida (Q05 or Q76.0), and encephalocele (Q01). All data were anonymized prior to analysis.

Incidence rates were calculated per 10,000 live births for each diagnosis and evaluated annually as well as in consecutive five-year intervals. Temporal trends in incidence were assessed using Poisson regression analysis. The proportion of prenatally diagnosed cases was evaluated using logistic regression models.

Sex distribution among affected cases was analyzed and compared with a control population consisting of children born in the same time periods without any registered congenital anomaly. Differences in sex distribution were assessed using Pearson’s chi-square test. Statistical analyses were performed using Stata software, version 15 (StataCorp LLC, College Station, TX, USA). A P value < 0.05 was considered statistically significant.

## Results

Between 1961 and 2020, a total of 2,521 cases of anencephaly, 3,391 cases of spina bifida, and 704 cases of encephalocele were recorded in the Czech Republic. Of the anencephaly cases, 1,456 occurred among live-born children, while 1,065 cases were diagnosed prenatally and the pregnancies were subsequently terminated. For spina bifida, 2,451 cases were recorded among live-born children and 940 cases were diagnosed prenatally. In encephalocele, 390 cases were identified among live-born children and 314 cases were prenatally diagnosed.

For the assessment of temporal trends, the study period was divided into twelve consecutive five-year intervals.

In anencephaly, the highest total incidence was observed in the first five-year period (1961– 1965), followed by a significant decline. During the subsequent seven five-year intervals up to the year 2000, incidence rates remained relatively stable, with no statistically significant differences between individual periods. A further decline was observed in the periods 2001– 2005 and 2006–2010, which showed the lowest incidence rates across the entire study period. In 2011–2015, a significant increase compared to the preceding decade was recorded. In the final period (2016–2020), total incidence decreased again compared to the previous five-year interval, although this change was not statistically significant. The proportion of prenatally diagnosed anencephaly cases increased markedly between 1985 and 1995 and subsequently fluctuated without significant variation between 94% and 98%. Incidence rates of anencephaly among live-born children and prenatally diagnosed cases are shown in Figure 1. The mean total incidence of anencephaly during the study period was 2.91 per 10,000 live births, with 0.14 among live-born children and 2.77 among prenatally diagnosed cases.

**Figure 1.**
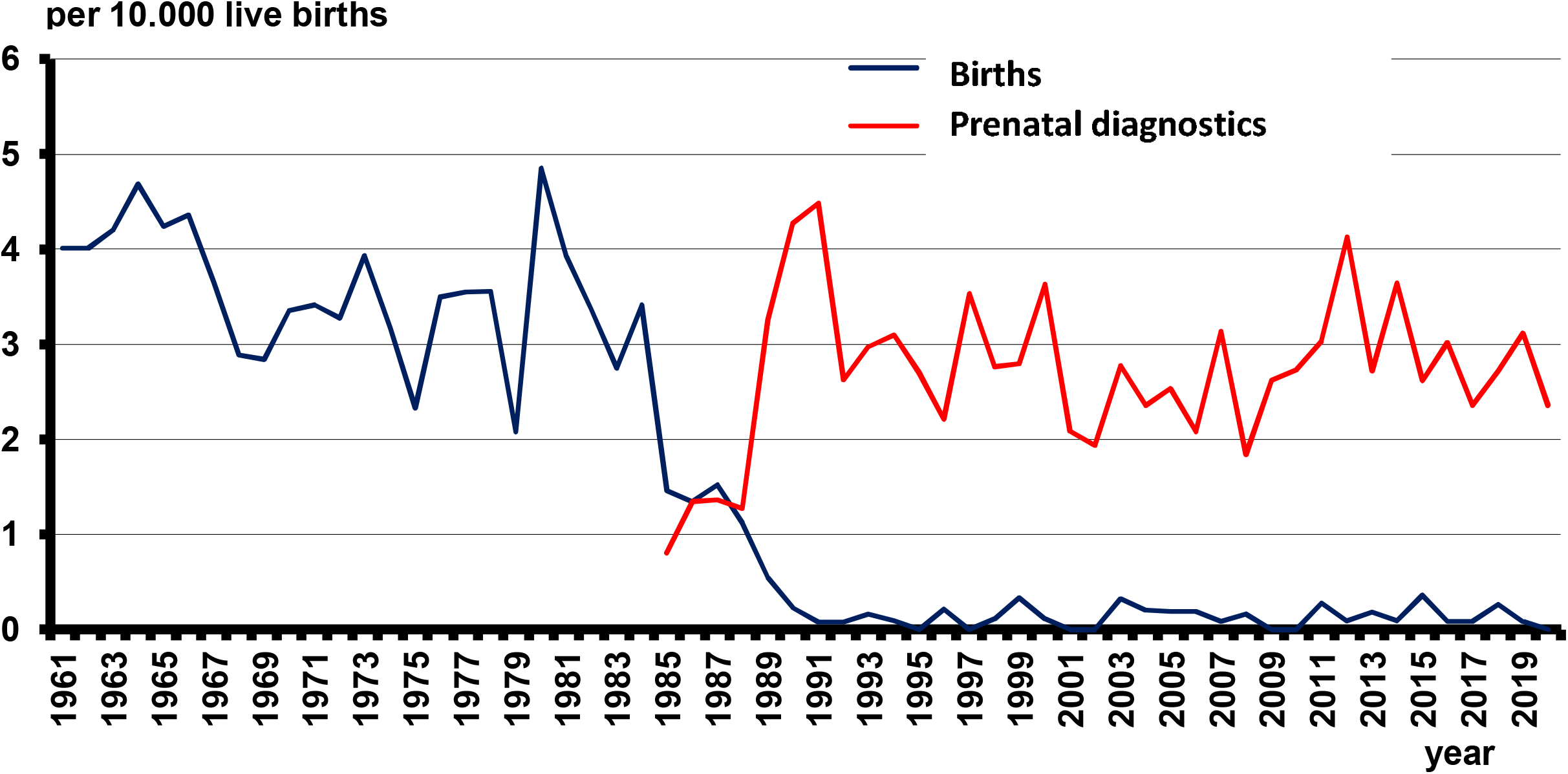
Anencephaly in the Czech Republic, 1961–2020: live births and prenatally diagnosed cases.

In spina bifida, the total incidence was significantly higher in the period 1961–1965 than in all subsequent five-year intervals. Over the following four periods up to 1985, incidence rates gradually declined without statistical significance, reaching the lowest value in 1981–1985. From that point onward until 2020, incidence rates fluctuated without a clear temporal trend. The proportion of prenatally diagnosed spina bifida cases increased continuously during the period 1986–2015. A significant increase was observed in 1991–1995 compared with 1986– 1990, followed by another significant increase in 1996–2000 compared with the preceding interval. In the final two five-year periods (2011–2020), the proportion of prenatally diagnosed cases was significantly higher than in any period prior to 2005. Incidence rates of spina bifida among live-born children and prenatally diagnosed cases are presented in Figure 2. The mean total incidence of spina bifida was 4.38 per 10,000 live births, with 1.54 among live-born children and 2.36 among prenatally diagnosed cases.

**Figure 2.**
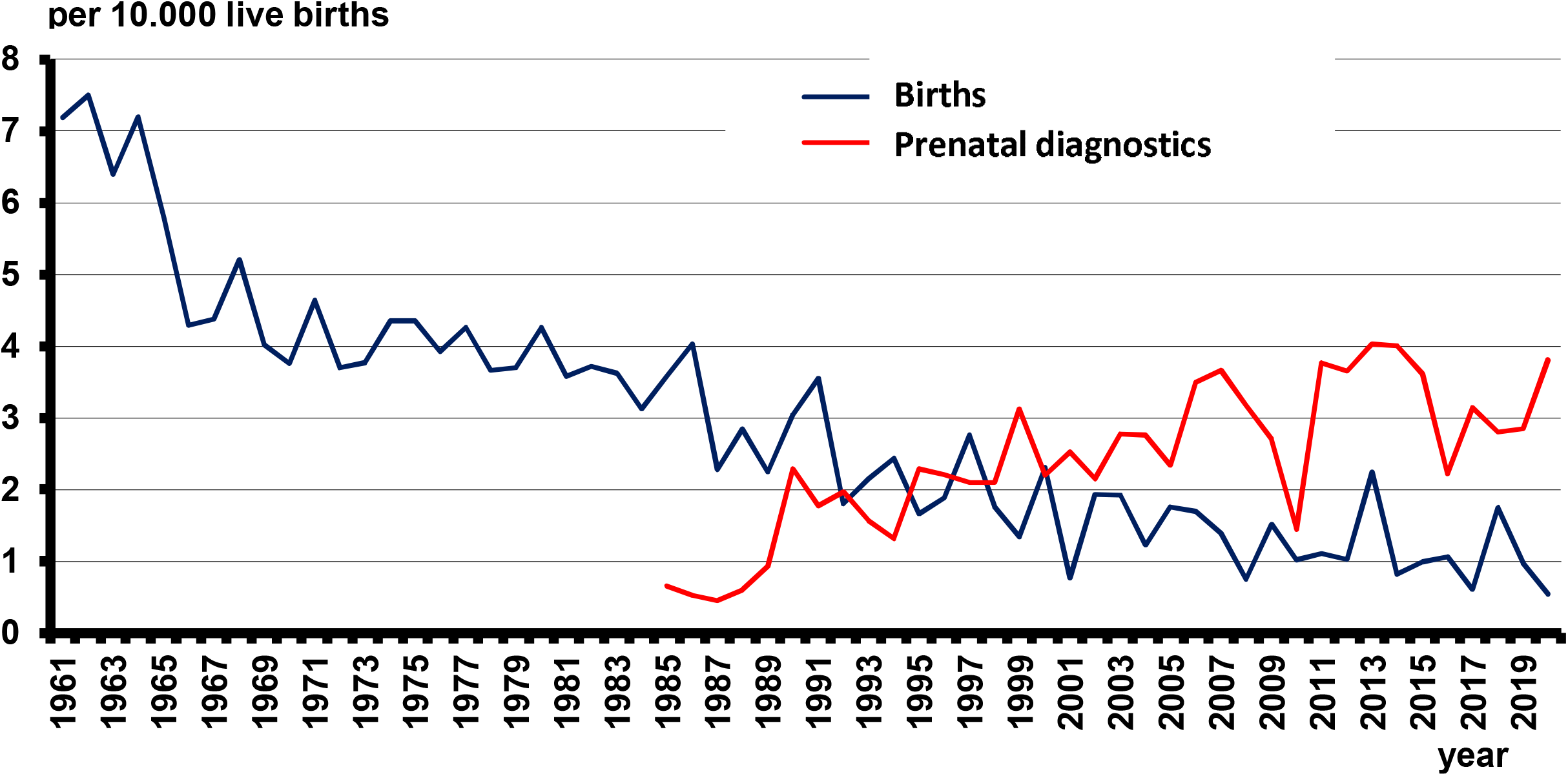
Spina bifida in the Czech Republic, 1961–2020: live births and prenatally diagnosed cases.

For encephalocele, 390 cases were recorded among live-born children and 314 cases were prenatally diagnosed. During the first 15 years of the study period (1961–1975), total incidence declined significantly, reaching its lowest level in 1971–1975, which differed significantly from all earlier and later periods. Subsequently, incidence rates showed an increasing trend, with values in 1976–1990 comparable to those observed in 1966–1970 and rates in 1991–2005 comparable to those in 1961–1965. In 2006–2010, incidence reached the highest level observed during the entire study period, significantly exceeding all previous intervals. In 2016–2020, incidence declined significantly compared with the preceding ten years, reaching levels similar to those observed in 2001–2005. The proportion of prenatally diagnosed encephalocele cases increased continuously during the first four five-year periods (1986–2005), although differences between these periods were not statistically significant. In the subsequent two periods (2006–2015), the proportion increased further and was significantly higher than in any period prior to 2005. In the final period (2016–2020), a non-significant decrease in the proportion of prenatally diagnosed cases was observed. Incidence rates of encephalocele among live-born children and prenatally diagnosed cases are shown in Figure 3. The mean total incidence of encephalocele during the study period was 1.24 per 10,000 live births, with 0.28 among live-born children and 0.96 among prenatally diagnosed cases.

**Figure 3.**
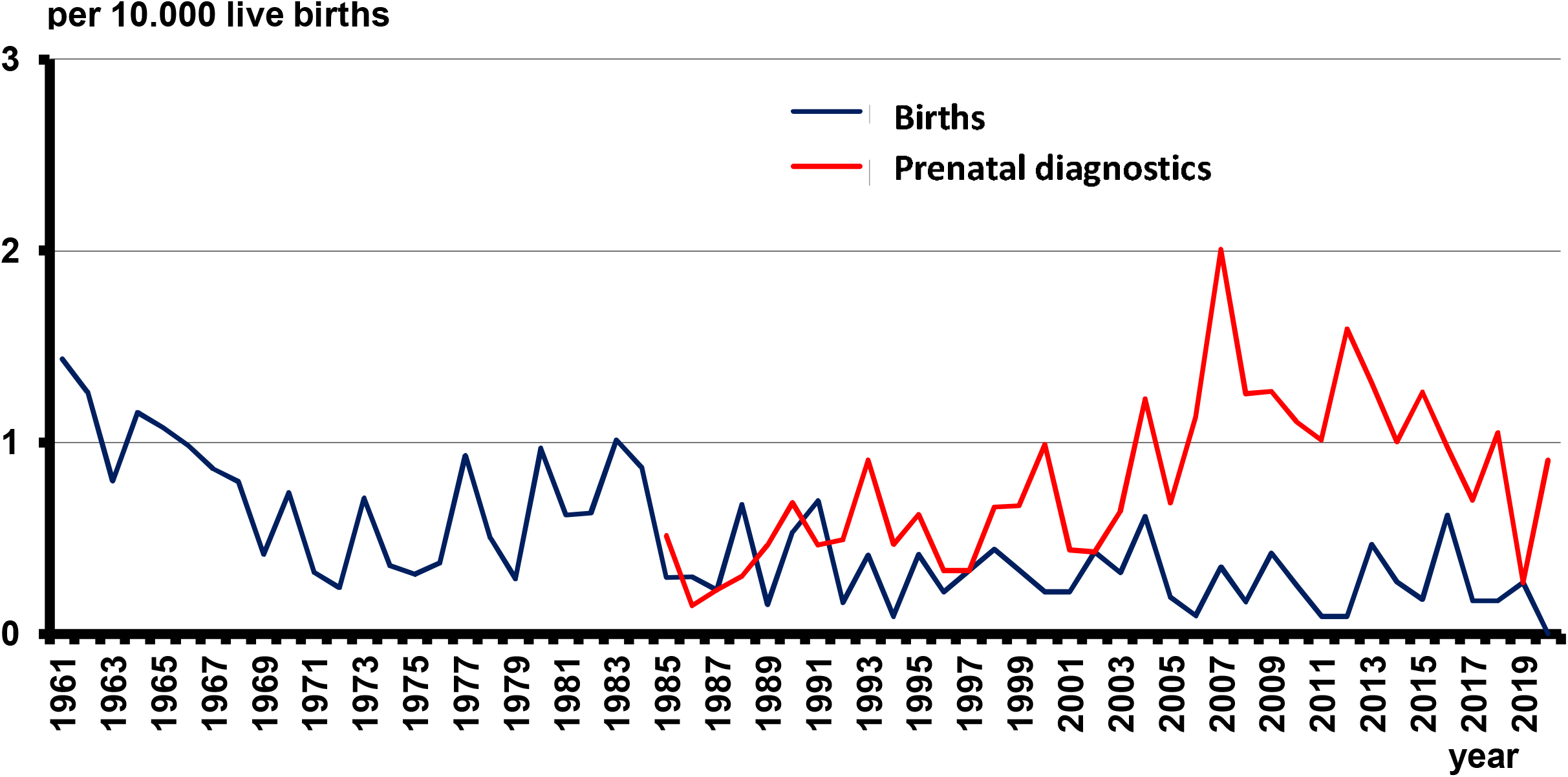
Encephalocele in the Czech Republic, 1961–2020: live births and prenatally diagnosed cases.

Sex distribution was analyzed across six consecutive ten-year periods and compared with a control population of children born without congenital anomalies.

In anencephaly, a higher proportion of females was observed among affected cases compared with the control population. Differences in the first three decades (1961–1990) were highly statistically significant (P < 0.001). In the subsequent three decades, the absolute number of live-born cases declined markedly, accompanied by a decrease in the proportion of affected females. In the period 1991–2000, a slightly higher proportion of affected males than females was observed. All differences in the second half of the study period were not statistically significant. The highest proportion of affected females (67.98%) was observed in 1971–1980, whereas the lowest proportion (45.45%) was recorded in 1991–2000. Sex distribution trends for anencephaly are presented in Figure 4.

**Figure 4.**
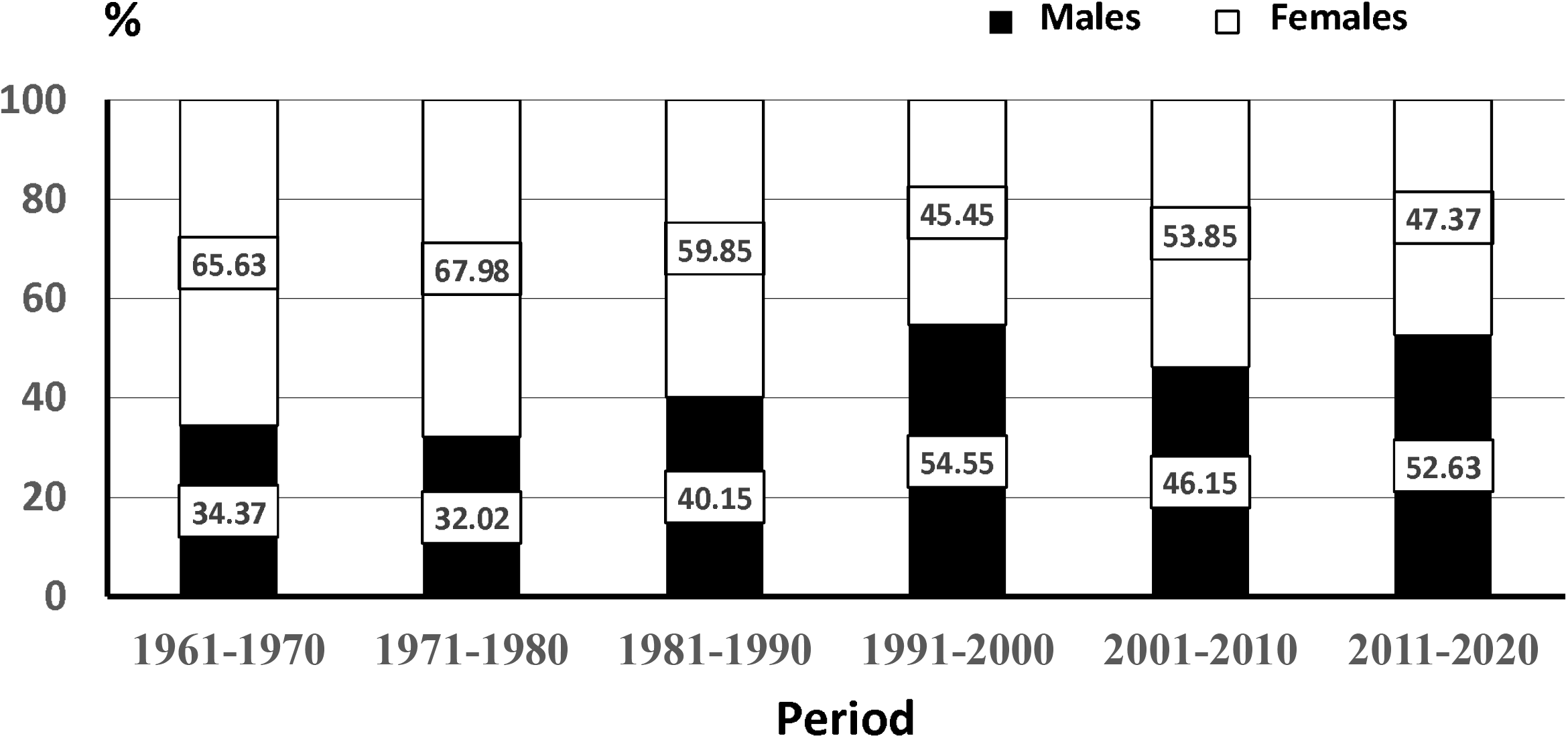
Anencephaly in the Czech Republic, 1961–2020: males and females.

In spina bifida, a higher proportion of affected females compared with males was observed in all six decades (Figure 5). In the first two decades (1961–1970 and 1971–1980), the proportion of females among affected cases was significantly higher than in the control population (P < 0.001). In the following three decades, female predominance persisted, although differences compared with the control population were not statistically significant. In the final decade (2011–2020), the proportion of affected females was the highest across all periods and, despite lower absolute case numbers, remained significantly different from the control population (P = 0.021). The highest proportion of affected females (59.83%) was observed in 2011–2020, while the lowest proportion (51.85%) occurred in 2001–2010.

**Figure 5.**
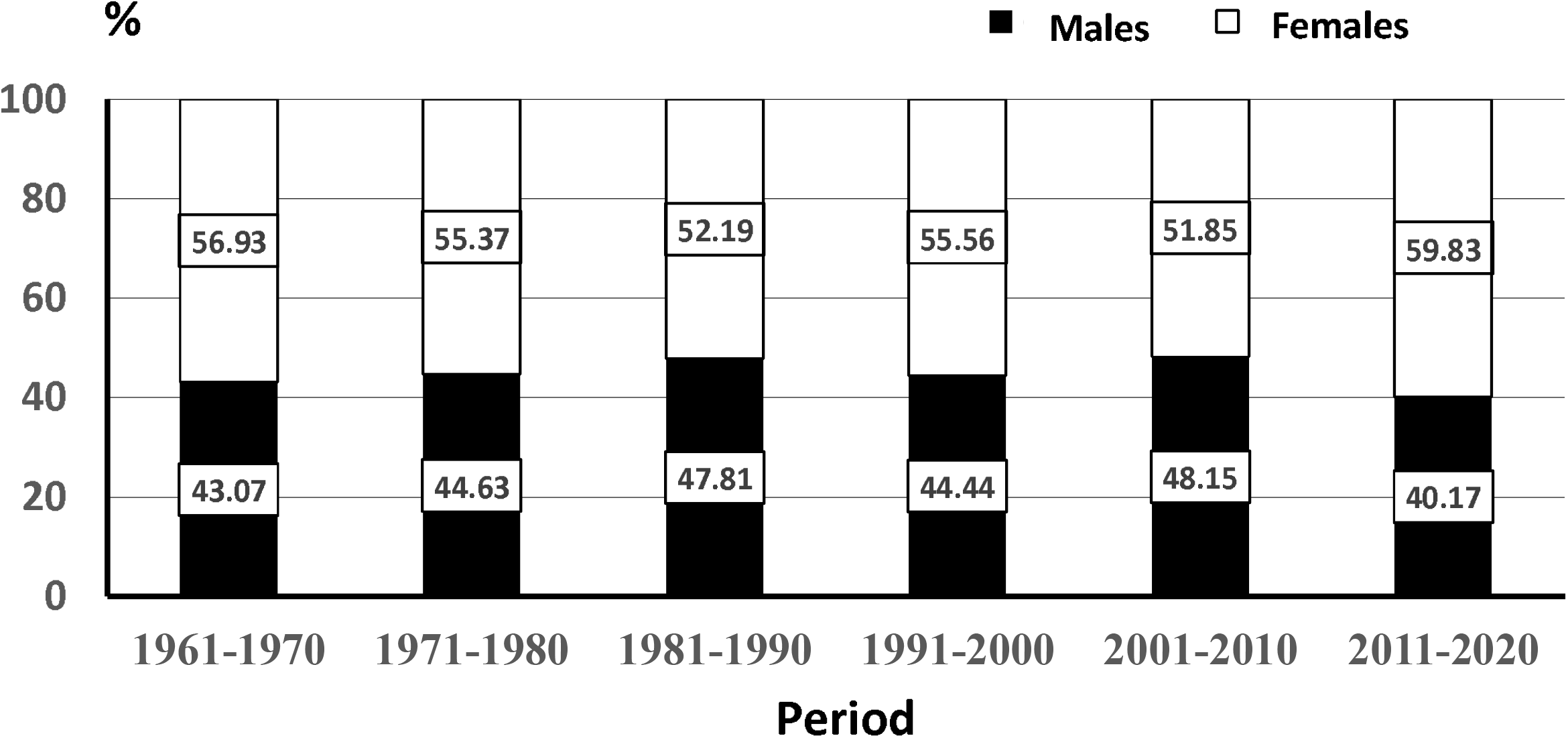
Spina bifida in the Czech Republic, 1961–2020: males and females.

Encephalocele was the least frequent diagnosis among the three NTDs analyzed (Figure 6). In the first two decades, a significantly higher proportion of females compared with the control population was observed. In 1981–1990, the proportion of affected females was similar to that observed two decades earlier (approximately 60%); despite reduced absolute case numbers, the difference compared with the control population remained statistically significant (P = 0.046). In 1991–2000, the proportion of affected females (64.3%) was higher than in 1961– 1970; however, due to small case numbers, the difference was not statistically significant. In the final two decades, a reversal of sex distribution was observed, with a higher proportion of affected males than females, although these differences were not statistically significant compared with the control population.

**Figure 6.**
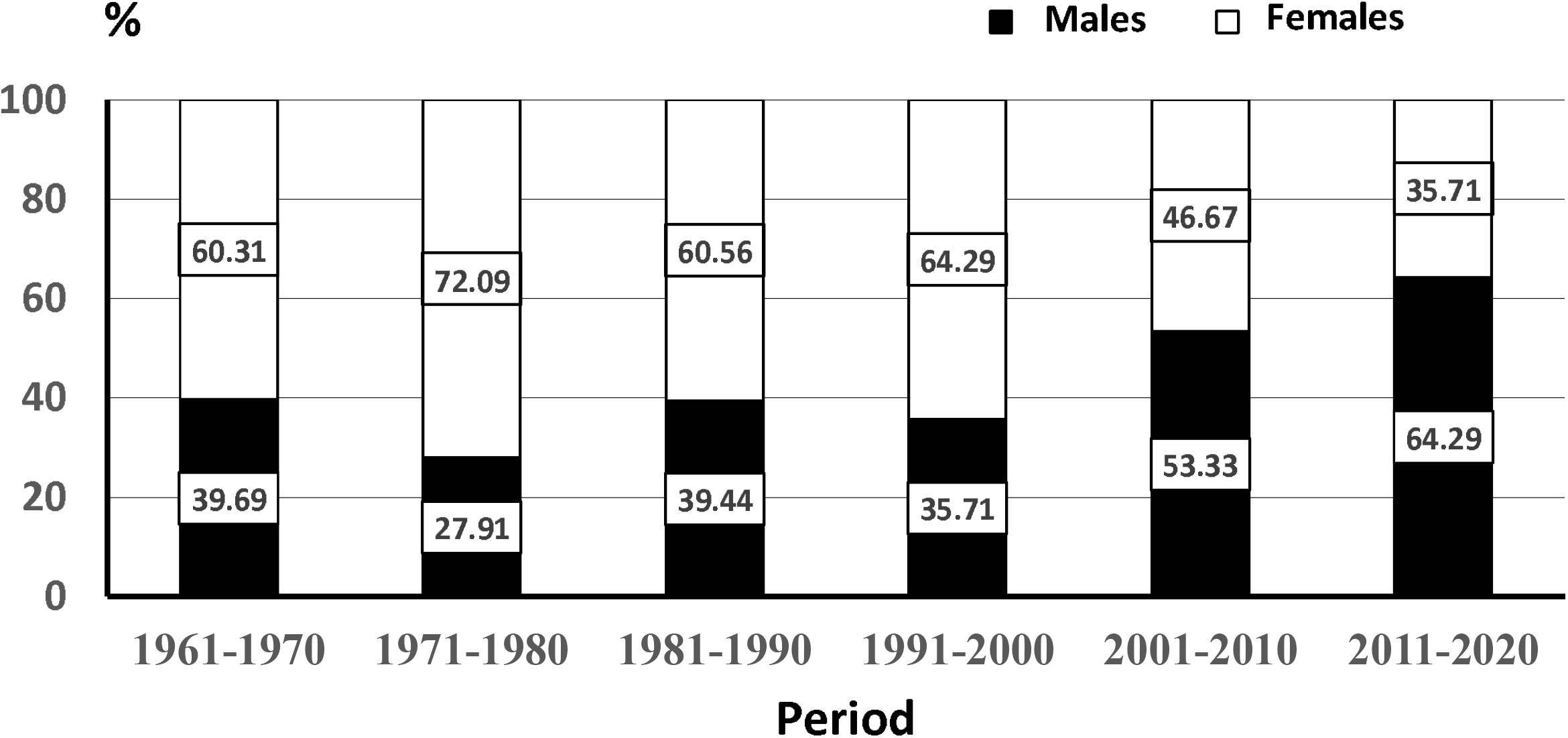
Encephalocele in the Czech Republic, 1961–2020: males and females.

## Discussion

This population-based study provides a comprehensive overview of long-term trends in incidence, prenatal diagnosis, and sex distribution of major neural tube defects in the Czech Republic over a 60-year period (Šípek et al., 2014). Our results confirm a substantial reduction in the incidence of anencephaly, spina bifida, and encephalocele among live-born children, which coincides with the increasing effectiveness and widespread implementation of prenatal diagnostic methods. However, despite this marked decline among live births, the overall population incidence of NTDs, including prenatally diagnosed and terminated pregnancies, has not shown a consistent decreasing trend (Šípek et al., 2002; Šípek et al., 2015).

These findings are in agreement with large European registry-based studies, including analyses from the EUROCAT network, which demonstrated that while the prevalence of live-born NTDs has declined, largely due to prenatal detection and termination of affected pregnancies, the total prevalence of NTDs has remained relatively stable over time (Khosnood et al., 2015). This pattern has been repeatedly attributed to the absence of mandatory folic acid fortification policies in most European countries, including the Czech Republic, despite long-standing recommendations for periconceptional folic acid supplementation (Šípek et al., 2013; Morris et al., 2021).

Temporal trends observed in our study further highlight differences among individual NTD subtypes. Anencephaly showed the highest incidence in the early 1960s, followed by a pronounced decline and subsequent stabilization, with fluctuations in later decades. Spina bifida exhibited a similar early decrease, followed by long-term fluctuation without a clear monotonic trend. In contrast, encephalocele demonstrated a more complex pattern, with an initial decline, a later increase peaking in the first decade of the 21st century, and a subsequent decrease (Morris et al. 2018). These heterogeneous trends suggest that individual NTD subtypes may be influenced by distinct combinations of genetic susceptibility, environmental exposures, and diagnostic practices (Knoshood et al., 2015; Morris et al., 2018).

A notable aspect of this study is the analysis of sex distribution over an extended time period. Consistent with previous reports, spina bifida showed a persistent predominance of affected females throughout the entire study period. In contrast, anencephaly and encephalocele exhibited a female predominance only in earlier decades, with a shift toward a higher proportion of affected males observed in recent years. Similar changes in sex ratios following improvements in folic acid intake or fortification have been reported in studies from South America and Asia, suggesting that sex-specific susceptibility to preventive measures may contribute to these observations (Lopez-Camelo et al., 2010; Poletta et al., 2018; Liu et al., 2018). Nevertheless, the biological mechanisms underlying sex differences in NTD occurrence remain poorly understood.

The main strength of this study lies in its long observation period and use of a nationwide registry with high population coverage. Limitations include potential changes in diagnostic criteria, reporting practices, and prenatal screening availability over time, which may have influenced observed trends. Additionally, individual-level data on maternal folic acid intake were not available, precluding direct assessment of its impact on incidence and sex distribution.

In conclusion, this long-term population-based analysis demonstrates that, in the Czech Republic, the incidence of major neural tube defects among live-born children has markedly declined over the past six decades, primarily as a result of increasingly effective prenatal diagnostics. However, the overall population incidence of these defects has not decreased, and in the case of encephalocele, a moderate increase was observed during part of the study period. While spina bifida consistently shows a predominance of affected females, the sex distribution of anencephaly and encephalocele has shifted in recent decades toward a higher proportion of affected males. In the absence of mandatory folic acid fortification, it remains uncertain to what extent voluntary periconceptional supplementation contributes to these changes. Continued population-based surveillance is therefore essential for monitoring long-term trends and evaluating preventive strategies for neural tube defects.

## Data Availability

Data availability statement:
The original individual data from official medical registries of the Czech Republic are not available for sharing due to national law. Aggregate datasets may be available upon request from the corresponding author.

## References

Akyol ME, Çelegen I, Basar I, Arabacı O. Hydrocephalus in encephalocele. Eur Rev Med Pharmacol Sci. 2022;26(15):5399–5405.

Atta CAM, Fiest KM, Frolkis ADF, et al. Global birth prevalence of spina bifida by folic acid fortification status: a systematic review and meta-analysis. Am J Public Health. 2016;106(1):e24–e34.

Avagliano L, Massa V, George TM, Qureshy S, Bulfamante GP, Finnell RH. Overview on neural tube defects: from development to physical characteristics. Birth Defects Res. 2019;111(19):1455–1467.

Brook FA, Estibeiro JP, Copp AJ. Female predisposition to cranial neural tube defects is not because of a difference between the sexes in the rate of embryonic growth or development during neurulation. J Med Genet. 1994;31(5):383–387.

Copp AJ, Adzick NS, Chitty LS, Fletcher JM, Holmbeck GN, Shaw GM. Spina bifida. Nat Rev Dis Primers. 2015;1:15007.

Czeizel AE, Dudás I. Prevention of first occurrence of neural tube defects with periconceptional vitamin supplementation. N Engl J Med. 1992;327:1832–1835.

Deak KL, Siegel DG, George TM, Gregory S, Ashley-Koch A, Speer MC; NTD Collaborative Group. Further evidence for a maternal genetic effect and a sex-influenced effect contributing to risk for human neural tube defects. Birth Defects Res A Clin Mol Teratol. 2008;82(10):662–669.

Hartill V, Szymanska K, Malik Sharif S, Wheway G, Johnson CA. Meckel–Gruber syndrome: an update on diagnosis, clinical management, and research advances. Front Pediatr. 2017;5:244.

Hassan AS, D. YL, Lee SY, Wang A, Farmer DL. Spina bifida: a review of the genetics, pathophysiology and emerging cellular therapies. J Dev Biol. 2022;10(2):22.

Iffy L. Thrice recurring anencephalus. Br J Clin Pract. 1963;17:83–84.

Khoshnood B, Loane M, de Walle H, et al. Long-term trends in prevalence of neural tube defects in Europe: population-based study. BMJ. 2015;351:h5949.

Liu J, Xie J, Li Z, et al. Sex differences in the prevalence of neural tube defects and preventive effects of folic acid supplementation among five counties in northern China. BMJ Open. 2018;8:e021476.

Lopez-Camelo JS, Castilla EE, Orioli IM. Folic acid flour fortification: impact on the frequencies of 52 congenital anomaly types in three South American countries. Am J Med Genet A. 2010;152A:2444–2458.

Morris JK, Addor MC, Ballardini E, et al. Prevention of neural tube defects in Europe: a public health failure. Front Pediatr. 2021;9:647038.

Poletta FA, Rittler M, Saleme C, et al. Neural tube defects: sex ratio changes after fortification with folic acid. PLoS One. 2018;13(3):e0193127.

Protzenko T, Dos Santos Gomes Junior SC, Bellas A, Salomão JFM. Hydrocephalus and occipital encephaloceles: presentation of a series and review of the literature. Childs Nerv Syst. 2021;37(11):3437–3445.

Record RG, McKeown T. Congenital malformations of the central nervous system. III. Risk of malformation in sibs of malformed individuals. Br J Prev Soc Med. 1950;4:217–220.

Salari N, Fatahi B, Fatahian R, Mohammadi P, Rahmani A, Darvishi N, et al. Global prevalence of congenital anencephaly: a systematic review and meta-analysis. Reprod Health. 2022;19(1):201.

Šípek A, Gregor V, Horáček J, Šípek A Jr, Klaschka J, Malý M. Prevalence of selected congenital anomalies in the Czech Republic: congenital anomalies of the central nervous system and gastrointestinal tract. Epidemiol Mikrobiol Imunol. 2015;64(1):47–53.

Šípek A, Gregor V, Horáček J, Šípek A Jr. National Registry of Congenital Anomalies of the Czech Republic: commemorating 50 years of the official registration. Cent Eur J Public Health. 2014;22(4):287–288.

Šípek A, Horáček J, Gregor V, Rychtaříková J, Dzúrová D, Mašátová D. Neural tube defects in the Czech Republic during 1961–1999: incidences, prenatal diagnosis and prevalences according to maternal age. J Obstet Gynaecol. 2002;22(5):501–507.

Šípek A Jr, Gregor V, Šípek A, Calda P. Primary prevention of congenital anomalies and the role of folic acid. Actual Gyn. 2013;5:47–51.

